# Colibactin genes are highly prevalent in the developing infant gut microbiome

**DOI:** 10.1101/2025.08.12.25333511

**Authors:** Shira Levy, Kathryn E McCauley, Rachel Strength, Emily S Robbins, Qing Chen, Sivaranjani Namasivyam, George Maxwell, Suchitra K Hourigan

**Affiliations:** Clinical Microbiome Unit, Laboratory of Host Immunity and Microbiome, Division of Intramural Research, National Institute of Allergy and Infectious Disease, National Institute of Health, Bethesda, MD; Bioinformatics and Computational Biosciences Branch, National Institute of Allergy and Infectious Diseases, National Institutes of Health, Bethesda, MD; Women’s Service Line, Inova Health System, Falls Church, VA, USA

**Keywords:** microbiota, neonate, neonatal intensive care unit, antibiotics, colorectal cancer

## Abstract

Early-life exposure to colibactin-producing *pks*+ gut bacteria is hypothesized to imprint mutations on the colorectal epithelium, increasing the risk of colorectal cancer later in life. We demonstrate an extremely high prevalence of *pks+* bacteria (>50% of infants) during the first two years of life, suggesting carriage is likely normal during early-life microbiome development. Further research is needed into the conditions in which carriage can lead to mutagenesis.

## Main

It is known that mutation signatures caused by colibactin, which is produced by bacteria carrying the *pks* pathogenicity island, contribute to the development of colorectal cancer (CRC)^1,2^. Recently it was found that these mutation signatures had higher mutation loads in countries with higher CRC incidence rates and were also enriched in early-onset CRC^3^. It was hypothesized that early-life exposure to colibactin-producing *pks*+ gut bacteria may cause imprinting of mutations on the colorectal epithelium, as *pks*+ bacteria was not found at the time of tumor sequencing^3^. While it is known that in the first few years of life gut microbiome development plays a critical role in immune system education, with disruptions in this critical window potentially having long lasting health impacts, the prevalence, dynamics and associated factors of *pks*+ gut bacteria carriage in early-life is unknown ^4-6^.

The first step in understanding the role colibactin may have in CRC development is to investigate the pks+ gut bacteria carriage in the early-life microbiome. We interrogated shotgun metagenomic sequencing of serial stool samples from the United States over the first two years of life from full term infants (N=55 infants, n=275 samples) and infants who were predominantly preterm and spent time in the neonatal intensive care unit (NICU) (N=128 infants, n=877 samples) (cohort characteristics in Supplementary Information). Using a previously established definition, where pks+ samples had at least one read in > 8 colibactin genes^3^, 31/55 (56%) of full term infants and 85/128 (66%) of NICU infants had at least 1 *pks*+ sample in the first two years of life. In the full-term cohort, the prevalence peaked at 6-12 months of age, with 38% of infants being positive, and then declined over time (Figure 1A). In the NICU infants, prevalence was significantly higher than full term infants at 2 months (OR = 3.02, P = 0.047), but also trended higher after 12 months (Figure 1A). Additionally, the limited NICU parental samples exhibited lower *pks*+ (4/18, 22%) and low levels of detection (mean of 11.5 RPM across all genes). In the one other study to our knowledge that looked at infant carriage of colibactin positive bacteria up to 1 month of life, high rates of *pks*+ (31%) were also seen^7^. Given the high prevalence of *pks*+ in the first two years of life, we hypothesize that carriage of colibactin producing bacteria may be part of the normal developing infant microbiome without adverse effects in most circumstances. Certainly, this is the case for toxigenic *Clostridioides difficile*, a potential carcinogenic bacterium and pathobiont, with very high prevalence rates in infancy without detrimental effects^8^. Indeed, in the full-term cohort where data was available, toxigenic *Clostridioides difficile* carriage was associated with significantly increased colibactin gene abundance (Generalized Estimating Equations (GEE) β = 1.30, P < 0.001).

**Figure 1.**
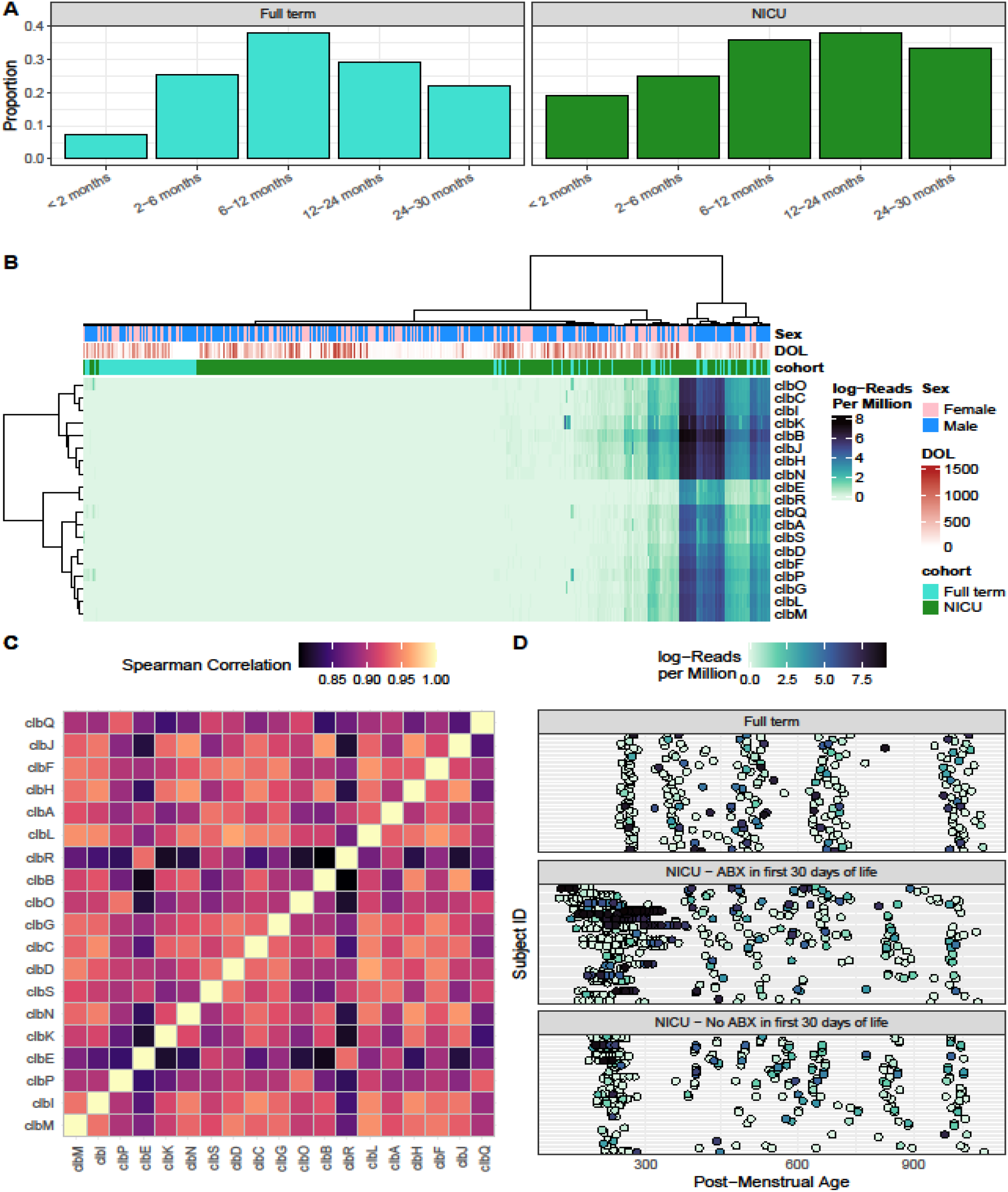
(A) Proportion of infants with pks+ samples. See supplemental methods for information on proportion calculations. (B) Heatmap of colibactin gene abundance as defined by log-transformed Reads per Million. Columns are annotated with infant sex (blue are males, and pink are females), Days of Life (light to dark red), and cohort (Full term in teal or NICU in green). (C) Heatmap of Spearman Correlations between each pair of colibactin genes; genes are arranged with Ward D2 clustering. (D) Colibactin gene abundance across samples; subjects are stratified into three groups based on cohort and antibiotic exposure; samples are arranged by post-menstrual age in days. Abbreviations: ABX: Antibiotics; DOL: Days of Life

We examined colibactin gene abundance, where colibactin genes were identified with at least one read in 491/1152 (43%) of samples from infants, with a mean of 342 reads per million (RPM) (Figure 1B). Individual colibactin genes were highly correlated with each other (Spearman Correlations > 0.8, Figure 1C), justifying utilizing the sum of all colibactin genes moving forward in further analyses. We next established longitudinal abundance of colibactin gene carrying bacteria (Figure 1D). A pattern was seen of very early-life carriage in NICU infants, predominantly in those who had received antibiotics in the first 30 days of life compared to the full-term cohort (Figure 1D, middle panel, GEE Day of Life by Cohort Interaction P = 0.0485).

When we examined the microbiome factors associated with colibactin abundance, no differences were seen in alpha diversity, however differences were seen in microbiome composition (Canberra distance, Figure 2A), in both the full-term (PERMANOVA R^2^ = 0.013, P = 0.001) and NICU (R^2^ = 0.012, P=0.001) cohorts. In the full-term cohort, *Escherichia* was the microbial genus associated with higher colibactin abundance while microbiota enriched with *Bifidobacterium, Bacteroides, Fecalibacterium* and *Blautia* exhibited limited colibactin abundance. In the NICU cohort, *Klebsiella* was the microbial genus associated with higher colibactin abundance while *Staphylococus* and *Enterococcus* enriched microbiomes exhibited limited colibactin abundance.

**Figure 2.**
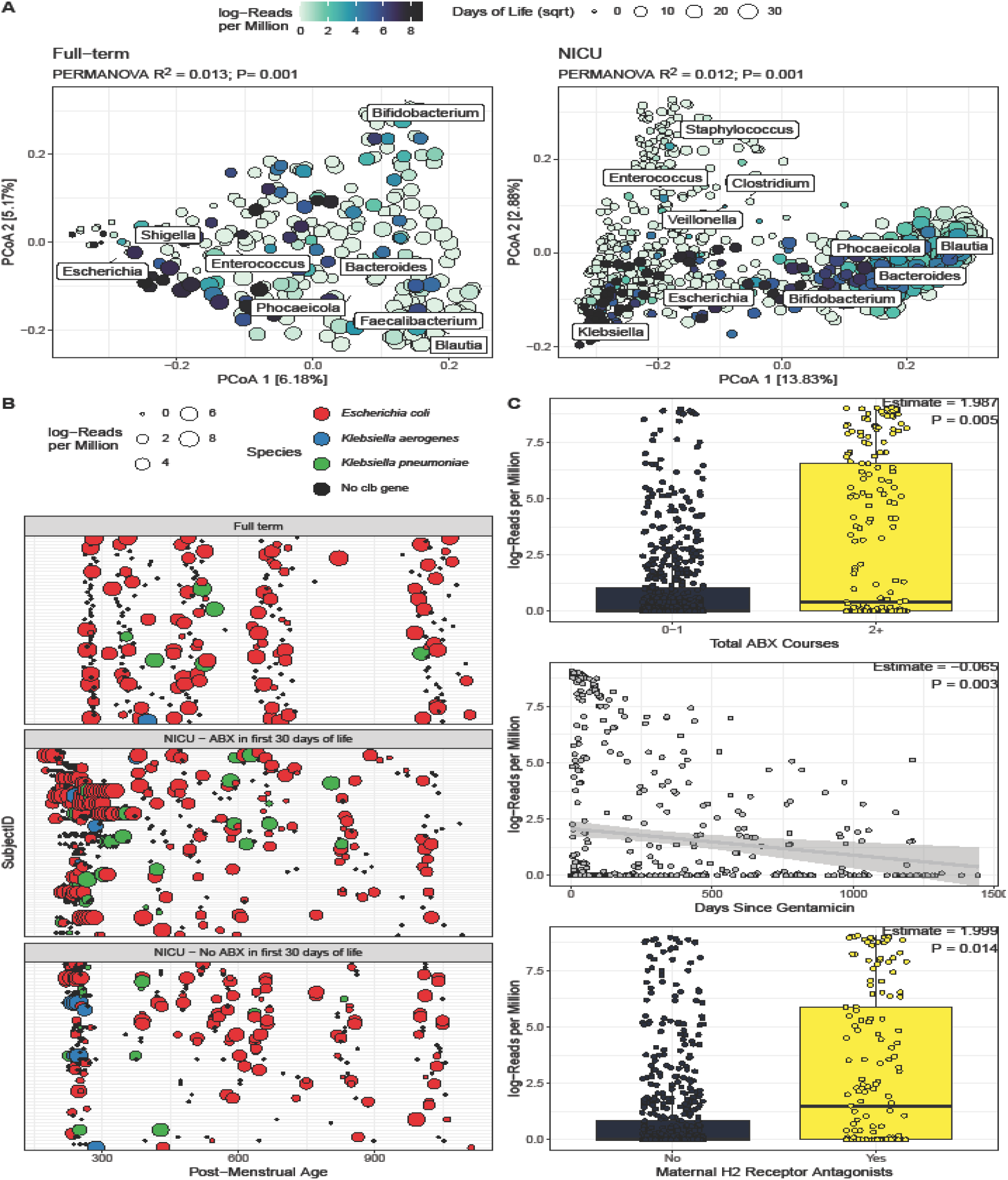
(A) Colibactin gene abundance compared to microbiome composition of Full term (left) and NICU cohorts (right). Points are colored by colibactin gene abundance and sized by Days of Life (square-root transformed). Biplot indicates the influence of the most dominant genera on community composition. (B) Bacterial host of colibactin genes across samples and time. Samples that did not have colibactin gene identified are represented in black; point size indicates the abundance of colibactin in the sample. Consistent with Figure 1C, samples are stratified by cohort and antibiotic exposure. (C) In the NICU, Colibactin abundance correlated with three variables: Total Antibiotics Courses, Days since Gentamicin, and Maternal H2 Receptor Antagonist Intake.

We then explored the specific species that were carrying colibactin genes and *Escherichia coli* was the dominant species in both cohorts (Figure 2B). However, infants in the NICU cohort who received antibiotics in the first 30 days of life had higher odds of having colibactin contained in *Klebsiella pneumoniae* compared to the full-term cohort (OR=3.49, P= 0.016). Of note, while most studies investigate the role of colibactin carrying *Escherichia coli* in CRC, colibactin carrying *Klebsiella pneumoniae* may also have a role in CRC progression^9^. We hypothesized that these genes may be sourced from the NICU environment, as we have previously shown colonization in infants from the NICU environment^10^. However, only 3/86 (3%) of environmental samples had more than 5 RPM for at least 8 colibactin genes, suggesting this was an unlikely source for these genes.

We next examined clinical factors associated with colibactin gene abundance. In the full-term cohort, after FDR correction, no clinical or demographic factors were significantly associated with colibactin gene abundance, including delivery mode, receiving breast milk and antibiotic use. In the NICU cohort, carriage in the first 6 months of life was positively associated with the total number of courses of antibiotics infants received within the NICU (P=0.001, Figure 2C), with the most common antibiotics received being gentamicin and ampicillin within the first 48 hours of life. Additionally, the days since receiving gentamicin negatively correlated with colibactin abundance (P<0.001, Figure 2C). Accordingly, systematic reviews have shown an increased risk of CRC with any antibiotic use and higher levels of antibiotic exposure^11,12^. A lack of association with antibiotic use in the full-term cohort may be due to the cohort consisting of generally healthy infants with minimal antibiotic use (Supplementary Information). In the NICU cohort, colibactin gene abundance was also higher in infants whose mothers had received H2 receptor antagonists during pregnancy (P=0.014, Figure 2C). While gastric acid suppressing mediations are known to alter the gut microbiome, theoretically making it more permissive to colibactin carrying bacteria, there is no direct evidence for this association or how mothers taking the medication affect the infants’ risk of carriage^12^.

Our study found that *pks*+ bacteria were highly prevalent (>50% of infants) in the developing early-life gut microbiome in the first two years of life. This far surpasses the rates of CRC, even in industrialized countries^3^. Given the very high prevalence, we hypothesize that this is likely a part of the normal developing gut microbiome, without harmful effects in most circumstances. Further exploration is warranted to assess when carriage of this bacteria in early-life leads to pathogenic colibactin expression. This may be achieved with lifespan cohort studies with serial sampling and genetic susceptibility determination, intestinal organoid models and transplant of infant stool into murine models. In addition, prevalence rates of *pks*+ bacteria carriage in early-life should be investigated in non-industrialized countries where CRC and early onset CRC rates are lower^3,13^. If conditions are found where early-life carriage of *pks*+ bacteria can cause imprinting of mutations associated with later CRC, modulation of the gut microbiome in early-life may represent a potential therapeutic target, given the microbiome compositional changes seen associated with carriage.

## Methods

### Subjects and Sample Collection

#### Full Term Cohort

Mothers were enrolled prenatally with informed consent in a longitudinal, prospective cohort study “The First 1000□Days of Life and Beyond” within the Inova Health System. All experimental protocols were approved by the Inova Health System and WCG Institutional Review Board (Inova protocol #15-1804, WCG protocol #20120204). In brief, serial stool samples were collected from infants at within the first 2 days of life (meconium) and approximately 2m, 6m, 12m, and 24m of age as previously described^14^. All samples were collected using previously validated methods and stored at −80°C until analysis^15^. Maternal demographic and clinical information including mode of delivery and use of prenatal antibiotics and peripartum antibiotics was collected via questionnaires and electronic medical records review. Infant data such as gestational age at delivery, use of antibiotics, and medical history were also collected from parents through questionnaires.

#### Neonatal Intensive Care Unit (NICU) cohort

Neonates who spent time in the Inova Fairfax Neonatal Intensive Care Unit (NICU) were enrolled in an observational longitudinal microbiome cohort study, “Neonatal Intestinal Microbiome: Impact on Infant and Early Childhood Health and Disease” as previously described^16^. The study was Institutional Review Board approved (WCG IRB 1300205) and parental informed consent was obtained. Neonates were enrolled within the first week of life and had an anticipated stay in the NICU of >5 days. While in the NICU stool samples were collected for microbiome analysis up to twice a week, and stored -80°C until analysis. Detailed demographic and clinical data was collected including delivery mode, gestational age, maternal peripartum antibiotic use and infant antibiotics.

After discharge from the NICU, follow-up surveys reporting health, illnesses, diet and a variety of exposures were collected approximately every 3–6 months until approximately 2 years of age (corrected for gestational age), accompanied by a stool sample collected by previously validated methods^17^. For this current study, neonates from the larger cohort were included if they had at least 1 stool sample from before 1 month of life, and at least 2 stool samples overall.

Environmental swabs were collected with a sterile cotton swab moistened with sterile saline and rolled over the area in a standardized manner from different areas in the NICU where the infants were hospitalized, including the wash sink at the nurses’ station, the desk at the nurses’ station, the light switch and the foam hand sanitizer pump as previously described^16,18^.

### DNA Extraction

DNA was extracted from fecal samples in two stages. First, approximately 50 mg of fecal material and 650 μL MBL lysis buffer from the PowerMicrobiome DNA/RNA EP Kit (Qiagen) were added to Lysis Matrix E (LME) tubes (MP Biomedicals). LME tubes were transferred to a Precelleys 24 Tissue Homogenizer (Bertin Technologies) and fecal samples were homogenized, centrifuged, with the resultant supernatant transferred to a deep-well 96-well plate. The second stage consisted of DNA isolation from the above supernatant using the MagAttract PowerMicrobiome DNA/RNA EP Kit (Qiagen) on an automated liquid handling system as detailed by the manufacturer (Eppendorf).

### Shotgun Metagenomic Sequencing

Total gene content of the microbiome was assessed through shotgun metagenomic sequencing. Metagenomic libraries were constructed from 100 ng of DNA as starting material using the Illumina DNA Prep kit. Illumina DNA/RNA UD Indexes were used to add sample-specific sequencing indices to both ends of the libraries. An Agilent 4200 TapeStation system with High Sensitivity D5000 ScreenTape (Agilent Technologies, Inc) was used to verify quality and assess final library size. A positive control (MSA-2002 20 Strain Even Mix Whole Cell Material (ATCC)) and a buffer extraction negative control were included. Metagenomic libraries were normalized and pooled at an equimolar concentration. Final pools were sequenced on the NovaSeq platformn using a paired-end (150×150) producing 2L×L150 bp paired-end reads (Illumina, Inc).

### Metagenomic Analysis

Extant metagenomic reads from the two cohorts underwent quality trimming with fastp and human reads were filtered with Kraken2 against the human genome using Whole Genome Shotgun Assembly (WGSA2) from the Nephele platform^19-21^. Environmental samples were trimmed with fastp and decontaminated for human reads with Ganon, as they were single-end and not amenable to the WGSA2 pipeline^22^. Reads from paired-end samples were assembled into contiguous sequences with metaSPAdes in Nephele, and genes were predicted using Prodigal^23,24^.

All non-human reads were separately aligned to the IHE3034 genome (RefSeq assembly: GCF_000025745.1) using bbmap, and reads per million for each of the 19 colibactin genes were obtained. In order to determine the bacteria containing colibactin, genes identified by Prodigal were aligned against the set of colibactin genes in the reference genome. Those aligning were taxonomically annotated with Ganon with a relative cutoff of 0.2^22^. The database contained NCBI complete genomes built June 25, 2025 with default parameters.

### Statistical Analysis

The prevalence of pks+ samples was calculated within age ranges used by visits for the Full term cohort; a 20-day buffer prior to the later sample collection timepoint was used to ensure participants early to their visit were grouped correctly. All samples from each time period were used; for each time period, the number of positive samples was divided by the total number of samples available for a subject to account for repeated sampling in the NICU cohort. These subject-level proportions were summed to create a weighted proportion at each timepoint. Permutational Analysis of Variance (PERMANOVA) was performed with adonis2 in the vegan package^25^. Correlations between colibactin genes were performed using Spearman correlation, and hierarchically clustered using the Ward D2 method. Generalized Estimating Equations (GEE) from the glmtoolbox package were used to compare longitudinal colibactin abundance with cohort factors, including *Clostridioides difficile* detection, using an Exchangeable correlation structure^26^.

## Supplemental Material

Supplemental Table 1: Description of demographic and clinical characteristics of the Full Term and NICU Cohorts.

## Acknowledgments

The content is solely the responsibility of the authors and does not necessarily represent the official views of the National Institutes of Health. The authors thank Kristy and Roger Crombie for their generous philanthropic donation toward this project in loving memory of their daughter Anna Charlotte. The authors thank the NIH Intramural Sequencing Center (NISC) for assistance with sequencing. This study used the Office of Cyber Infrastructure and Computational Biology (OCICB) High Performance Computing (HPC) cluster at the National Institute of Allergy and Infectious Diseases (NIAID), Bethesda, MD.

## Ethics approval and consent to participate

All subjects provided signed informed consent prior to collection and storage of samples. This study was Institutional Review Board approved (WCG IRB 1300205 and WCG protocol 20120204). Consent forms included language authorizing publication of findings.

## Data Availability

Data are publicly available from the Sequence Read Archive for the full-term cohort under the accession <u>PRJNA988496</u>, and for the NICU cohort under the accession <u>PRJNA1280936</u>. Environmental samples are available from <u>PRJNA417283</u>.

This paper does not report original code.

## Author Contributions

Conceptualization: SL, SKH

Methodology: SL, KEM, RS, ESR, QC, SN, GM, SKH

Investigation: SL, KEM, RS, ESR, QC, SN, GM, SKH

Data Curation: SL, KEM

Visualization: SL, KEM

Funding acquisition: SKH

Project administration: SL, SKH

Supervision: SKH

Writing – original draft: SL, KEM, SKH

Writing – review & editing: SL, KEM, RS, ESR, QC, SN, GM, SKH

## References

1 Chen, B. et al. Contribution of pks(+) E. coli mutations to colorectal carcinogenesis. Nat Commun 14, 7827 (2023). 10.1038/s41467-023-43329-5

2 Pleguezuelos-Manzano, C. et al. Mutational signature in colorectal cancer caused by genotoxic pks(+) E. coli. Nature 580, 269–273 (2020). 10.1038/s41586-020-2080-8

3 Díaz-Gay, M. et al. Geographic and age variations in mutational processes in colorectal cancer. Nature (2025). 10.1038/s41586-025-09025-8

4 Gensollen, T., Iyer, S. S., Kasper, D. L. & Blumberg, R. S. How colonization by microbiota in early life shapes the immune system. Science 352, 539–544 (2016). 10.1126/science.aad9378

5 Yatsunenko, T. et al. Human gut microbiome viewed across age and geography. Nature 486, 222–227 (2012). 10.1038/nature11053

6 Cox, L. M. et al. Altering the intestinal microbiota during a critical developmental window has lasting metabolic consequences. Cell 158, 705–721 (2014). 10.1016/j.cell.2014.05.052

7 Tsunematsu, Y. et al. Mother-to-infant transmission of the carcinogenic colibactin-producing bacteria. BMC Microbiol 21, 235 (2021). 10.1186/s12866-021-02292-1

8 Mani, J. et al. Epidemiological and microbiome associations of Clostridioides difficile carriage in infancy and early childhood. Gut Microbes 15, 2203969 (2023). 10.1080/19490976.2023.2203969

9 Kaur, C. P. et al. Presence of Polyketide Synthase (PKS) Gene and Counterpart Virulence Determinants in Klebsiella pneumoniae Strains Enhances Colorectal Cancer Progression In-Vitro. Microorganisms 11 (2023). 10.3390/microorganisms11020443

10 Chaudhary, P. P. et al. Vaginal delivery provides skin colonization resistance from environmental microbes in the NICU. Clin Transl Med 13, e1506 (2023). 10.1002/ctm2.1506

11 Liu, Y. C. et al. Effects of antibiotic exposure on risks of colorectal tumors: a systematic review and meta-analysis. J Transl Med 23, 682 (2025). 10.1186/s12967-025-06727-5

12 Aneke-Nash, C., Yoon, G., Du, M. & Liang, P. Antibiotic use and colorectal neoplasia: a systematic review and meta-analysis. BMJ Open Gastroenterol 8 (2021). 10.1136/bmjgast-2021-000601

13 Mäklin, T. et al. Geographical variation in the incidence of colorectal cancer and urinary tract cancer is associated with population exposure to colibactin-producing Escherichia coli. Lancet Microbe 6, 101015 (2025). 10.1016/j.lanmic.2024.101015

14 Subramanian, P. et al. Delivery Mode Impacts Gut Bacteriophage Colonization During Infancy. Gut Microbes Reports 2, 2464631 (2025). 10.1080/29933935.2025.2464631

15 McDonald, D. et al. American Gut: an Open Platform for Citizen Science Microbiome Research. mSystems 3 (2018). 10.1128/mSystems.00031-18

16 Hourigan, S. K. et al. Comparison of Infant Gut and Skin Microbiota, Resistome and Virulome Between Neonatal Intensive Care Unit (NICU) Environments. Front Microbiol 9, 1361 (2018). 10.3389/fmicb.2018.01361

17 Wong, W. S. W. et al. Collection of non-meconium stool on fecal occult blood cards is an effective method for fecal microbiota studies in infants. Microbiome 5, 114 (2017). 10.1186/s40168-017-0333-z

18 Chaudhary, P. P. et al. Shotgun metagenomic sequencing on skin microbiome indicates dysbiosis exists prior to the onset of atopic dermatitis. Allergy 78, 2724–2731 (2023). 10.1111/all.15806

19 Chen, S., Zhou, Y., Chen, Y. & Gu, J. fastp: an ultra-fast all-in-one FASTQ preprocessor. Bioinformatics 34, i884–i890 (2018). 10.1093/bioinformatics/bty560

20 Wood, D. E., Lu, J. & Langmead, B. Improved metagenomic analysis with Kraken 2. Genome Biol 20, 257 (2019). 10.1186/s13059-019-1891-0

21 Weber, N. et al. Nephele: a cloud platform for simplified, standardized and reproducible microbiome data analysis. Bioinformatics 34, 1411–1413 (2018). 10.1093/bioinformatics/btx617

22 Piro, V. C., Dadi, T. H., Seiler, E., Reinert, K. & Renard, B. Y. ganon: precise metagenomics classification against large and up-to-date sets of reference sequences. Bioinformatics 36, i12–i20 (2020). 10.1093/bioinformatics/btaa458

23 Nurk, S., Meleshko, D., Korobeynikov, A. & Pevzner, P. A. metaSPAdes: a new versatile metagenomic assembler. Genome Res 27, 824–834 (2017). 10.1101/gr.213959.116

24 Hyatt, D. et al. Prodigal: prokaryotic gene recognition and translation initiation site identification. BMC Bioinformatics 11, 119 (2010). 10.1186/1471-2105-11-119

25 Oksanen J B. F., Kindt R, Legendre P, Minchin P, O’Hara B, Simpson G, Solymos P, Stevens H, Wagner H. Vegan: Community Ecology Package. (2022).

26 <https://CRAN.R-project.org/package=glmtoolbox > (

