## Supplementary Information for "Colibactin genes are highly prevalent in the developing infant gut microbiome"

Supplemental Table 1: Description of demographic and clinical characteristics of the Full Term and NICU Cohorts.

| **Infant Characteristics** | **Full Term Cohort (n=55)** | **NICU Cohort (n=128)** |
| --- | --- | --- |
| **Sex (n, %)** | | |
| Male | 28 (51%) | 76 (59%) |
| Female | 27 (49%) | 52 (41%) |
| **Ethnicity (n, %)** | | |
| Not Hispanic or Latino | 52 (95%) | 90 (70%) |
| Hispanic or Latino | 3 (5%) | 13 (10%) |
| Unknown | 0 (0%) | 25 (20%) |
| **Race (n, %)** | | |
| White or Caucasian | 38 (69%) | 65 (51%) |
| Black or African American | 6 (11%) | 17 (13%) |
| Asian | 7 (13%) | 21 (16%) |
| More than one Race | 3 (5%) | 3 (2%) |
| Other | 1 (2%) | 8 (6%) |
| Unknown | 0 (0%) | 14 (11%) |
| **Mode of Delivery** | | |
| Vaginal Delivery | 33 (60%) | 26 (20%) |
| Cesarean Section | 22 (40%) | 102 (80%) |
| **Gestational Age (n, %)** | | |
| Extremely Preterm (<28 weeks) | 0 (0%) | 21 (16%) |
| Very Preterm (28-<32 weeks) | 0 (0%) | 35 (27%) |
| Moderate Preterm (32-<34 weeks) | 0 (0%) | 28 (22%) |
| Late Preterm (34-<37 weeks) | 0 (0%) | 31 (24%) |
| Term (≥37 weeks) | 55 (100%) | 13 (10%) |
| **Infant Antibiotics in the first 30 days of life (n, %)** | | |
| Yes | 1 (2%) | 73 (57%) |
| No | 54 (98%) | 55 (43%) |
| **Infant antibiotics prior to 2 months (n, %)** | | |
| Yes | 7 (13%) | 74 (58%) |
| No | 48 (87%) | 54 (42%) |
| **Infant antibiotics between 2 to 6 months (n, %)** | | |
| Yes | 6 (11%) | Not Available |
| No | 49 (89%) | Not Available |
| Unknown | 0 (0%) | Not Available |
| **Infant antibiotics between 6 to 12 months (n, %)** | | |
| Yes | 17 (31%) | 24 (19%) |
| No | 38 (69%) | 64 (50%) |
| Unknown | Not applicable | 40 (31%) |
| **Infant antibiotics between 12 to 24 months (n, %)** | | |
| Yes | 8 (15%) | 33 (26%) |
| No | 47 (85%) | 67 (52%) |
| Unknown | 0 (0%) | 28 (22%) |
| **Any Breast Milk before 2 months (n, %)** | | |
| Yes | Not Available | 105 (82%) |
| No | Not Available | 23 (18%) |
| **Any Breast Milk at 6 months (n, %)** | | |
| Yes | 40 (73%) | Not Available |
| No | 9 (16%) | Not Available |
| Unknown | 6 (11%) | Not Available |

| **Maternal Characteristics** | **Full Term Cohort (n=55)** | **NICU Cohort (n=128)** |
| --- | --- | --- |
| **Maternal Age in years, mean (±SD)** | 34.03 (±4.13) | 33.85 (±5.65) |
| **Maternal Pre-Pregnancy BMI (kg/m^2^), mean (±SD)** | 24.23 (±3.94) | 25.46 (±6.44) |
| **Maternal prenatal H2 receptor antagonists (n, %)** | | |
| Yes | 0 (0%) | 17 (13%) |
| No | 0 (0%) | 99 (77%) |
| Unknown | 0 (0%) | 12 (9%) |
| **Maternal prenatal antibiotics (n, %)** | | |
| Yes | 7 (13%) | 34 (27%) |
| No | 48 (87%) | 94 (73%) |
| **Maternal peripartum antibiotics (n, %)** | | |
| Yes | 29 (53%) | 123 (96%) |
| No | 26 (47%) | 5 (4%) |
